# Improving Doctor-Patient Communication Using Large Language Models - Results from an Experimental Study

**DOI:** 10.1101/2025.10.23.25338067

**Authors:** Christian Böffel, Stella J. Soyka, Sibu Mundi, Wolfgang Wick, Sabine J. Schlittmeier, Varun Venkataramani

## Abstract

**Importance:** Medical jargon poses significant barriers to patient comprehension of healthcare information, potentially affecting treatment adherence and health outcomes.

**Objective:** To investigate whether large language models (LLMs) can improve patient understanding of medical notes by translating complex medical terminology into comprehensible lay language.

**Design, Setting, and Participants:** This experimental online study was conducted between August 27 and October 8 2024 using a within-subjects design. Participants were recruited from a university population and included 63 adults aged 19-30 years (52 female [82.5%]; mean age 21.9 years), who completed the full study protocol.

**Interventions:** Four fictional medical notes representing typical neurological cases (stroke, neuromyelitis optica, meningitis, subarachnoid hemorrhage) were created by neurologists and translated into lay language using GPT-4. Each participant viewed two original and two translated notes in counterbalanced order.

**Main Outcomes and Measures:** Primary outcomes included objective comprehension measured through content-related questions, self-paced reading times and subjective ratings of understanding, difficulty, empathy, and mental effort on 5-point Likert scales.

**Results:** The translated notes demonstrated significant improvements across all measured dimensions, except for average reading times. Participants achieved higher comprehension scores (effect size details to be added), reported greater subjective understanding, perceived higher empathy, and experienced reduced mental effort and negative emotions when reading translated versus original medical notes.

**Conclusions and Relevance:** LLM translation of medical notes significantly improved both objective and subjective patient understanding. From a psychological perspective, these results align with predictions from cognitive load theory, emphasising the importance of adapting language complexity to reduce processing demands in communication between laypeople and experts. These findings suggest potential for integrating AI-assisted communication tools in clinical practice to enhance patient comprehension and engagement, though implementation considerations including accuracy validation and clinician workflow integration require further investigation.

## Introduction

Effective communication between healthcare providers and patients represents a cornerstone of quality medical care, directly influencing patient satisfaction, treatment adherence, and health outcomes.^1–3^ However, the pervasive use of specialized medical terminology in healthcare settings creates substantial barriers to patient comprehension, potentially compromising the quality of care and patient safety.

Health literacy, defined as the capacity to obtain, process, and understand basic health information needed to make appropriate health decisions,^4^ remains problematically low across diverse patient populations. The National Assessment of Adult Literacy found that only 12% of adults have proficient health literacy skills, while 36% have basic or below-basic health literacy.^5^ This deficit becomes particularly pronounced when patients encounter medical documentation filled with clinical jargon, abbreviations, and complex terminology.

Medical notes, while primarily designed for communication between healthcare professionals, often contain crucial information for patients regarding their diagnosis, treatment plans, and follow-up care. However, patients frequently struggle to comprehend these documents, leading to several concerning outcomes. Studies demonstrate that patients commonly misinterpret medical terminology, with “positive” test results or “outstanding” imaging findings often perceived as favorable news despite indicating pathological conditions.^2^ Additionally, patients frequently fail to recognize medical jargon terms as specialized vocabulary, potentially leading to incomplete understanding of their health status.^3^ Such examples illustrate that even functional health literacy, the first of three levels in Nutbeam’s (2000) model, is often insufficient. It refers to basic skills in reading and understanding medical information. If patients struggle at this foundational level, it becomes even more unlikely that they will be able to engage in interactive communication regarding their health (level 2) or critically reflect on medical content (level 3).

The prevalence of abbreviations in medical records further compounds comprehension difficulties. Grossman Liu et al. demonstrated that expanding abbreviations and acronyms significantly improved patient understanding of their health records, suggesting that even minor modifications to medical language can yield meaningful improvements in comprehension.^6^ These communication barriers disproportionately affect patients with lower educational attainment, with Lehmann et al. observing that highly educated patients were less likely to report physician use of confusing language.^7^

The implications of inadequate patient understanding extend far beyond momentary confusion. Miscomprehension of medical information can lead to medication non-adherence, delayed seeking of appropriate care, inappropriate self-management behaviors, and increased anxiety and distress.^8–10^ In worst-case scenarios, misunderstanding critical medical information can result in preventable adverse outcomes, emergency department visits, and healthcare complications.

Research in health communication consistently demonstrates that patients who better understand their medical conditions exhibit improved treatment adherence, better self-management behaviors, and enhanced health outcomes.^11–13^ Conversely, communication failures contribute to medical errors, with estimates suggesting that communication problems play a role in approximately 70% of adverse events in healthcare settings.^14^ From a psychological perspective, these comprehension difficulties can be interpreted as resulting from excessive intrinsic and extraneous cognitive load, as described by cognitive load theory^15^. Intrinsic load stems from the inherent complexity of medical concepts, while extraneous load stems from the unfamiliar linguistic form in which they are presented. Therefore, reducing linguistic complexity without compromising informational integrity may lower processing demands and facilitate understanding and engagement.

### Facilitating Layman-Expert Communication Using Artificial Intelligence

Recent advances in large language models (LLMs) present unprecedented opportunities to address the persistent challenge of medical communication complexity. These models demonstrate remarkable capabilities in natural language processing, including the ability to adjust language complexity while preserving semantic meaning, which is particularly relevant for medical translation applications.

LLMs excel at producing natural, comprehensible text with freely varying degrees of complexity, making them well-suited for translating medical jargon into patient-friendly language. Preliminary evidence suggests considerable potential for improving patient understanding of medical records through AI-assisted translation.^16^ However, rigorous empirical evaluation of these tools remains limited.

A small-scale pilot study by Bala et al. used AI software to translate medical notes into plain language, yielding promising results despite lacking statistical power for definitive conclusions.^16^ Separately, a systematic review regarding AI chatbots in healthcare contexts generally demonstrates positive outcomes in terms of user satisfaction and usability.^17^

Notably, LLM-generated medical content may offer advantages beyond improved comprehension. Ayers et al. found that licensed medical professionals rated AI-generated responses to patient questions more favorably than physician responses in terms of both quality and empathy,^18^ suggesting that AI tools might enhance multiple dimensions of patient-provider communication. At the same time, participants are less likely to follow medical advice, if they believe that an AI was involved in creating it.^19^

### Study Objectives and Hypotheses

This study investigates the extent to which large language models can improve the information interface between doctors and patients by translating medical vocabulary and contexts into layman’s terms. Given the potential consequences of communication failures in healthcare, this research addresses a critical gap in our understanding of AI-assisted medical communication tools.

#### Primary Objectives

- To assess the viability of LLMs as mediators of patient-doctor communication
- To evaluate whether patient understanding of medical information can be improved using LLM-generated language simplifications
- To assess the accuracy and applicability of LLM translations of medical content

#### Secondary Objectives

- To identify factors influencing comprehension of translated medical content
- To explore potential implementation barriers and considerations for clinical practice

#### Hypotheses

1. Translated medical notes will lead to significantly higher frequency of correct answers to comprehension questions compared to original medical notes.
2. Translated medical notes will lead to significantly higher subjective ratings of understanding and perceived empathy, and reduced mental effort, confusion, negative emotions and reading times compared to original notes.

## Methodology

An experimental online study was conducted using Limesurvey^20^ to compare the comprehension and perception of LLM-translated medical notes by laypeople against the original versions created by neurologists. First, the preparation and translation of the medical notes is described before reporting the experimental study.

### Participants

Participants were recruited through a university mailing list and could receive partial course credit as compensation. A total of 76 participants took part in the experiment, 63 participants completed the experiment and were included in the data analysis. Of these, 52 (82.5%) self-identified as female, 9 as male (14.3%), and 2 as non-binary (3.2%). The age range was 19-30 years (*M* = 21.9, *SD* = 2.1). All participants gave informed consent before participating in this study. The medical knowledge of the participants was assessed using a three-level item and 9.5% stated that they had no medical knowledge, 87.3% stated that they had limited medical knowledge, e.g. from a related field of study such as psychology. Only 3.2% stated that they had extensive medical knowledge, e.g. from medical training or a dedicated medical degree. Participants were compensated with partial course credit.

For the central hypothesis test, the within-subject comparison between the original and simplified tests, a sample of at least 45 participants was aimed for to achieve a statistical power of (1-β) = 0.95 for medium-size effects *d*_z_ = 0.5 and an α-level of 0.05 in a one-sided t-test for dependent samples. This calculation is based on G*Power 3.1.9.7.^21^

### Study Design

The study used a within-subjects design with the conditions “original” vs. “translation”. For each condition, two medical notes were shown per participant for a total of four medical notes per participant. Each of the four notes featured a different fictional patient with a different diagnosis and history. Which medical note was shown as original or translation was counterbalanced across participants, and the order of the medical notes was randomized.

### Material Development

#### Medical Note Creation

Four fictional medical notes were developed to represent typical cases encountered in neurological practice. All notes and the corresponding translations were created in German. Each note described a different patient scenario featuring one of four common neurological diagnoses: acute ischemic stroke, neuromyelitis optica spectrum disorder, bacterial meningitis, and subarachnoid hemorrhage.

The medical notes were crafted to reflect authentic clinical documentation, incorporating typical medical terminology, diagnostic procedures, treatment recommendations, and follow-up instructions commonly found in neurological practice. Each note was structured to include patient presentation, diagnostic workup, clinical findings, treatment plan, and discharge instructions.

#### Expert Validation

Three neurologists independently reviewed the doctor’s notes for accuracy. Only minor inaccuracies were found in the GPT-translated version, none of which affected the overall understanding of the letter or introduced critical misinformation. GPT consistently failed to explain certain drugs and medications, although this was outside the scope of the prompt.

#### LLM Translation Process

Original medical notes were translated into lay language using GPT-4 (OpenAI, accessed 06/2024). The translation process aimed to preserve all essential medical information while replacing technical terminology with patient-comprehensible language, expanding abbreviations, and providing explanatory context where appropriate. The prompts, the medical notes and their translation can be found under: https://github.com/Varaman/CLARITY_LLM

#### Translation Validation

All translated versions underwent review by the same three neurologists who validated the original notes. This review process ensured that translations maintained medical accuracy while successfully simplifying language complexity.

#### Comprehension Assessment

A total of eight items were used to assess how participants subjectively perceived the medical notes. Participants indicated their agreement on a 5-point scale (1= “do not agree at all” to 5 “completely agree”) to items querying their subjective understanding, the perceived mental effort, confusion, anger and empathy. The items and the predicted effects are shown in Table 1.

**Table 1.**
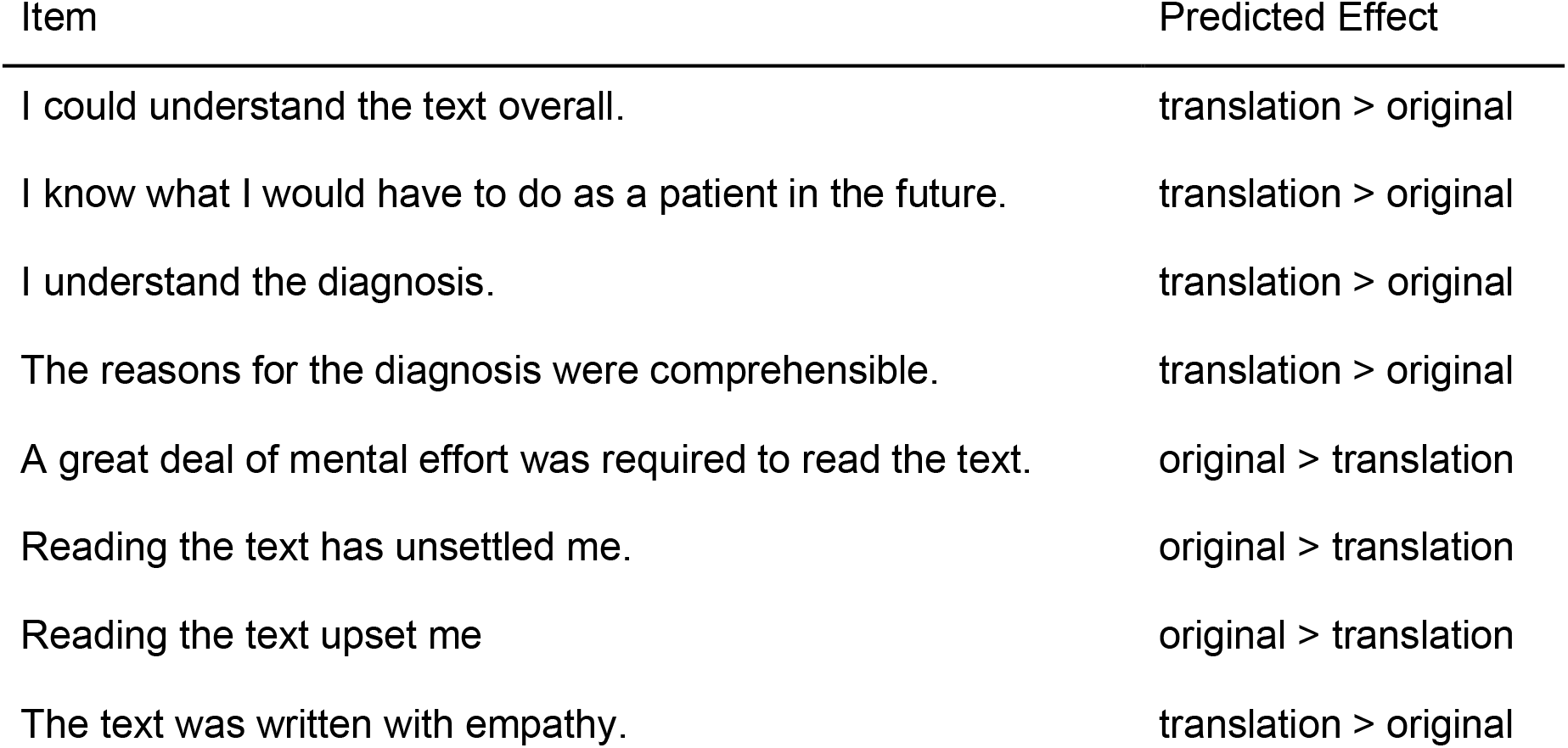
Predicted Effects of Used Items.

### Procedure

The study was conducted using the online survey platform Limesurvey^20^ and the participants accessed the study via a participation link. Participants first received information about the study and gave informed consent. Participants were then shown four medical notes with a maximum viewing time of 10 minutes each. When the timer expired or the participant decided to stop reading, the reading time was recorded, and the comprehension and perception questions were asked. This was followed by questions about age, gender, level of education, employment status and general medical knowledge. Finally, participants were given the opportunity to provide any additional comments about this study, and the study concluded with a short debriefing.

### Data Analysis

First, 13 participants who did not complete the experiment were excluded from the data analysis. Then it was checked whether the comprehension questions were answered correctly, and the relative frequency of correct answers was calculated for each medical note. The mean frequency of correct answers and the means of each subjective rating item were then calculated for the two conditions: original vs. translation. Additionally, mean reading times in seconds for each condition were calculated. With these means, repeated measures t-tests were performed to check for significant differences between the originals and the translations.

## Results

The results of the repeated measures t-tests comparing original medical notes to their translation are shown in Table 2.

**Table 2.** Results of two-tailed, repeated measures t-tests comparing original and translated doctor’s letters (original-translation), Bonferroni-adjusted for multiple comparisons.

| Variable/Item | Original |  | Translation |  | *t*(62) | *p* | Cohen's *d* |
| --- | --- | --- | --- | --- | --- | --- | --- |
|  | *M* | *SD* | *M* | *SD* |  |  |  |
| % Correct | **61%** | 14% | **71%** | 14% | -4.89 | < .001 | -0.62 |
| Comprehensible | **2.33** | 0.80 | **3.83** | 0.67 | -11.67 | < .001 | -1.47 |
| Clear instructions | **2.83** | 0.98 | **3.83** | 0.84 | -7.47 | < .001 | -0.94 |
| Clear diagnosis | **2.46** | 0.96 | **3.73** | 0.79 | -9.56 | < .001 | -1.20 |
| Clear d. rationale | **2.66** | 0.93 | **3.71** | 0.73 | -8.77 | < .001 | -1.11 |
| Mental Effort | **4.31** | 0.59 | **3.54** | 0.95 | 5.80 | < .001 | 0.73 |
| Caused uncertainty | **3.32** | 1.03 | **2.34** | 0.99 | 8.98 | < .001 | 1.13 |
| Caused anger | **2.42** | 1.06 | **1.62** | 0.72 | 6.59 | < .001 | 0.83 |
| Empathically written | **1.90** | 0.78 | **2.85** | 1.05 | -7.29 | < .001 | -0.92 |
| Mean reading time (s) | **258** | 110 | **291** | 112 | -4.00 | .002 | -0.50 |

*Note.* "% Correct" refers to the proportion of correctly answered single-choice comprehension-questions with two to four choice alternatives.

The tests revealed significant advantages of the translation with medium to very large effect sizes for all dependent variables measured, except for average reading times, where significantly shorter reading times were observed for the original versions compared to the translations.

### Further Analyses of Reading Times and Word Count

We conducted additional analyses to investigate the unexpected, faster reading times for the original compared to the translated texts. First, when comparing the original texts to their translations, it is evident that the translation process increases word count, e.g., by adding additional explanations or writing out abbreviations in full. As a result, the average word-count of the translated texts is 35.65% higher compared to the original (*M*_*original*_ = 653.5; *M*_*Translation*_ = 886.5). In comparison, the observed increase in average reading time from 258 to 291 seconds, only constitutes a relative increase of 12.79%.

Furthermore, a closer look at the reading times at the individual and per trial level reveals reading times that cast doubt on whether the texts could have been read in such a short time. To establish the fastest plausible reading time, a value of 300 wpm (words per minute) was chosen based on the study by Radner et al., 2002^22^. This is a conservative estimate, as the sentences in Radner et al. were simple and a reading speed of 300 words per minute was achieved by very few individuals, with lower values being much more likely (cf., 230±28 wpm for university students). Applying this to originals and translations separately, a cut-off for plausible reading times of 130s for originals and 177s for translations was obtained. When comparing individual trials in both conditions, 16 original trials and 19 translation trials were identified as implausible out of a total of 252 trials. A X^2^-Test revealed that this did not represent a significant deviation from an equal distribution (X^2^ = 0.257, *p* = .61).

### Exploratory Analyses

Pearson’s correlations were calculated to explore the relationship between the measured variables. This section describes some important observations based on these, and the complete results of the correlation analysis are shown in Table 3. Regarding the 4 items on perceived comprehensibility and clarity of the medical notes, we observed a significant correlation between the individual items, which is to be expected. More importantly, perceived comprehensibility was negatively associated with negative emotions after reading the notes, as measured by items 7 and 8. Regarding the perceived empathy of the texts, higher perceived empathy was associated with perceived comprehensibility, lower perceived uncertainty and longer average reading times and most of these correlations are descriptively more pronounced in the translation compared to the original. Additionally, longer average reading times were associated with higher perceived comprehensibility and empathy.

**Table 3.** Pearson-Correlations for Study Variables. *Above the diagonal: Translation; below the diagonal: Original*.

| Variable | 1 | 2 | 3 | 4 | 5 | 6 | 7 | 8 | 9 | 10 |
| --- | --- | --- | --- | --- | --- | --- | --- | --- | --- | --- |
| 1. % Correct | — | .30* | .29* | .37** | .32* | -.28* | -.39** | -.21 | .20 | .20 |
| 2. Comprehensible | .22 | — | .37** | .76** | .39** | -.64** | -.47** | -.25 | .58** | .27* |
| 3. Clear instructions | .27* | .44** | — | .40** | .36** | -.14 | -.45** | -.31* | .38** | .22 |
| 4. Clear diagnosis | .28* | .64** | .66** | — | .58** | -.47** | -.48** | -.24 | .53** | .28* |
| 5. Clear d. rationale | .29* | .45** | .56** | .65** | — | -.163 | -.39** | -.18 | .45** | .24 |
| 6. Mental Effort | -.09 | -.35** | -.09 | -.16 | -.22 | — | .43** | .24 | -.31* | -.15 |
| 7. Caused uncertainty | -.29* | -.39** | -.32* | -.31* | -.28* | .27* | — | .40** | -.28* | -.05 |
| 8. Caused anger | -.39** | -.47** | -.34** | -.42** | -.41** | .04 | .45** | — | -.21 | -.07 |
| 9. Empathically written | .12 | .37** | .24 | .38** | .32* | -.17 | -.27* | -.19 | — | .29* |
| 10. Mean reading time | .15 | .12 | .35** | .24 | .35** | -.02 | -.06 | -.15 | .26* | — |

\**p* < .05. \*\**p* < .01.

## Discussion

This study provides robust empirical evidence supporting the potential of large language models to potentially improve patient understanding of medical information. The results demonstrate clear advantages of LLM-translated medical notes across both objective comprehension measures and subjective evaluation dimensions, with effect sizes indicating clinically meaningful improvements.

### Primary Findings and Clinical Implications

The significant improvement in comprehension scores for translated notes directly supports our primary hypothesis and aligns with growing evidence and consensus that reducing linguistic complexity enhances understanding of health information.^23,24^ These findings could have implications for clinical practice, suggesting that AI-assisted translation tools could serve as valuable intermediaries in patient-provider communication.

The comprehensive benefits observed across subjective measures, including enhanced perceived understanding, reduced mental effort, decreased negative emotions, and increased perceived empathy, indicate that LLM translation addresses multiple barriers to effective medical communication simultaneously. Psychologically, these benefits can be interpreted through the lens of cognitive load theory: By simplifying linguistic form while preserving semantic content, LLMs likely reduce extraneous load, allowing more cognitive resources for schema acquisition and integrative comprehension. The finding that translated notes were perceived as more empathetic is particularly noteworthy, as empathetic communication is strongly associated with improved patient satisfaction, treatment adherence, and health outcomes.^25–27^ The increase in reading time was unexpected at first but can largely be explained by the increased text length resulting from the translation. While this may appear to be a downside at first, exploratory analyses revealed beneficial correlations between reading times and other variables that could suggest higher levels of engagement.

### Implications for Healthcare Practice and Health Equity

The implementation of LLM-assisted medical translation could have profound implications for addressing health disparities related to health literacy. Given that limited health literacy disproportionately affects vulnerable populations including older adults, individuals with lower educational attainment, and racial/ethnic minorities,^28–30^ accessible communication tools could help reduce these disparities.

Healthcare systems could integrate LLM translation capabilities into electronic health record systems, allowing automatic generation of patient-friendly versions of medical notes, discharge summaries, and treatment plans. This approach could be particularly valuable in settings with high patient volume or limited time for individualized explanation, such as emergency departments or busy primary care practices.

The enhanced comprehension demonstrated in this study could translate to improved treatment adherence, better self-management behaviors, and reduced healthcare utilization through prevention of misunderstanding-related complications. The economic implications of improved patient understanding, such as reduced readmissions, fewer medication errors, and enhanced preventive care uptake, warrant further investigation.

### Implementation Considerations and Challenges

Despite the promising results, several considerations must be addressed before widespread clinical implementation. First, ensuring translation accuracy and medical safety requires robust validation processes. While this study included expert review of translations, scalable implementation would necessitate automated accuracy verification systems or streamlined clinician review protocols.

Second, integration into clinical workflows must balance efficiency gains with quality assurance. The additional physician time required for review and approval of AI-generated translations must be weighed against potential benefits in reduced patient queries, improved adherence, and decreased downstream complications.

Legal and regulatory considerations present additional implementation challenges. Healthcare organizations must establish clear policies regarding AI-generated patient communications, including liability frameworks, quality assurance protocols, and patient disclosure requirements about AI involvement in communication materials.

### Trust and Acceptance in AI-Mediated Healthcare Communication

Patient acceptance of AI-generated medical information represents a critical implementation factor. While research suggests patients may be initially hesitant to follow AI-generated medical advice compared to physician recommendations,^29^ the context of physician-authored content translated by AI may face less resistance, particularly given the established acceptance of AI in translation applications.

Healthcare providers must also be prepared to address patient questions about AI involvement in their care communications. Transparent disclosure policies and education about AI translation capabilities and limitations will be essential for maintaining patient trust and therapeutic relationships.

### Study Limitations and Future Research Directions

Several limitations should be considered when interpreting these results. The study population consisted primarily of young, educated, female participants with some, but limited medical knowledge, potentially limiting generalizability to broader patient populations. Additionally, patients are likely to acquire some degree of medical knowledge about their own medical condition. The personal relevance of medical notes is expected to be higher for real patients than for the study population. Future research should examine LLM translation effectiveness across diverse age groups, educational backgrounds, and health literacy levels.

The use of fictional medical notes, while ensuring standardization and ethical compliance, may not fully capture the complexity and variability of real clinical documentation. Studies using authentic (de-identified) medical records would provide more ecologically valid assessments of translation effectiveness.

Although the task required participants to briefly retain medical information in their memory, the relatively short-term nature of comprehension assessment does not address retention of medical information over a longer period of time. Longitudinal studies examining whether improved initial understanding translates to sustained knowledge retention and appropriate health behaviors would strengthen the evidence base for clinical implementation.

Future research should also investigate optimal prompting strategies for medical translation, comparative effectiveness of different LLM models, and integration approaches that maximize clinical utility while minimizing physician burden.

### Technological and Methodological Considerations

The reading time findings, while initially counterintuitive, provide valuable insights into patient engagement with medical information. The disproportionate increase in reading time relative to word count suggests that participants may have been more willing to engage with translated materials, or that the simplified language facilitated more thorough comprehension processes.

The identification of implausibly short reading times in both conditions highlights the challenges of online study methodologies and the importance of attention checks in health communication research. Future studies might benefit from more stringent engagement monitoring or alternative assessment approaches.

### Broader Context and Future Directions

This research contributes to the growing body of evidence supporting AI applications in healthcare communication. The findings align with broader trends toward patient-centered care and shared decision-making, providing technological solutions to persistent communication challenges.

Future investigations should examine the effectiveness of LLM translation for other medical specialties, different types of medical documents (laboratory results, imaging reports, treatment protocols), and various clinical populations. Additionally, research into optimal integration strategies, cost-effectiveness analyses, and long-term patient outcomes will be crucial for evidence-based implementation decisions.

The potential for personalized translation approaches, adapting language complexity based on individual users’ prior knowledge and cognitive capacity medical history, or stated preferences, represents an exciting frontier for AI-assisted healthcare communication that warrants further exploration.

## Conclusions

This study demonstrates that large language model translation of medical notes significantly improves both objective comprehension and subjective patient experience across multiple dimensions. The findings provide strong empirical support for the potential role of AI-assisted communication tools in enhancing patient understanding and engagement with medical information.

The comprehensive benefits observed, including improved comprehension, reduced cognitive burden, decreased negative emotions, and enhanced perceived empathy, suggest that LLM translation addresses fundamental barriers to effective patient-provider communication. These improvements could contribute to better health outcomes through enhanced patient understanding, improved treatment adherence, and more informed healthcare decision-making.

However, successful clinical implementation requires careful attention to accuracy validation, workflow integration, regulatory compliance, and patient acceptance considerations. Future research should focus on real-world implementation studies, long-term outcome assessments, and optimization of AI-assisted communication tools for diverse clinical settings and patient populations.

The integration of LLM technology into healthcare communication represents a promising avenue for addressing persistent health literacy challenges and advancing patient-centered care. As AI capabilities continue to evolve, the potential for more sophisticated, personalized, and effective patient communication tools will likely expand, offering new opportunities to bridge the communication gap between healthcare providers and the patients they serve.

## Data Availability

All data produced in the present study are available upon reasonable request to the authors.

https://github.com/Varaman/CLARITY_LLM

## Competing Interest Statement

The authors have no competing interest to disclose.

## Funding Statement

This research received no external funding.

## Data Availability Statement

All data produced in the present study are available upon reasonable request to the authors.

## Ethics Statement

The Ethics Committee of the Medical Faculty of Heidelberg University gave ethical approval for this work (S-658/2023).

## Acknowledgments

We thank Fabienne Willnecker for her valuable assistance in creating the experiment.

## Notes

### Competing Interest Statement

The authors have declared no competing interest.

### Author Declarations

The Ethics Committee of the Medical Faculty of Heidelberg University gave ethical approval for this study (S-658/2023).

